# Spread of the plague in Venice, 1630–1631: epidemic entropy in a “natural experiment”

**DOI:** 10.1101/2025.10.06.25335371

**Authors:** Jonathan L. Hwang, Ariktha Srivathsan, Michael S. Deiner, Seth Blumberg, Travis C. Porco, Thomas M. Lietman

## Abstract

Precise modeling of epidemic spread is difficult. One explanation is that disease spread is inherently stochastic. This would suggest that the distribution of cases across geographic regions would progress towards that more favored by chance. If the epidemic proceeds long enough, the allocation of cases could approach that most expected, maximizing Boltzmann–Gibbs–Shannon entropy. Here, we tested these hypotheses on mortality data from the Venetian 1630–1631 plague epidemic. Entropy per case (intensive) of the quantile function (distribution of parishes ranked by case rates) increased from an effective number of 7.32 parishes (95% CI 3.32–12.55 parishes) to 47.9 parishes (47.5–48.9 parishes) out of 50 total, indicating that the quantile function approached a uniform maximum entropy distribution. Intensive entropy of the probability density function (parishes categorized by cumulative case rate) increased from 0.63 nats (0.32–0.93 nats) to 1.75 nats (1.53–1.87 nats). The PDF approached a Gaussian distribution. The Kullback–Leibler divergence decreased from 0.84 nats (0.71–1.42 nats) to 0.12 nats (0.083– 0.35 nats). These findings quantify how disease spreads and demonstrate that observed heterogeneity in infections between regions may in some circumstances be explained by chance alone.

## Introduction

Forecasting the time course and geographical extent of epidemics often fails.^1–6^ However, predicting the distribution of cases across communities may be achievable.^7–12^ If disease transmission were in large part driven by stochasticity, the accuracy of small-scale predictions would be limited, while large-scale behavior would be predictable. In particular, observed case distributions (macrostates) with more possible combinatorial arrangements (microstates) would be favored over time. Therefore, the distribution of infections across communities would have increasing Boltzmann–Gibbs–Shannon entropy over time. Furthermore, if the epidemic persisted long enough, it is plausible that this distribution would approach the distribution with the greatest number of such arrangements, a maximum entropy distribution. Specific maximum entropy distributions, in particular, the uniform, exponential, or Gaussian distributions, can be characterized by a small number of constraints (0, 1, and 2 sufficient parameters, respectively). Beyond these constraints, maximum entropy distributions make minimal assumptions to model the distribution of infections across geographic regions during an epidemic.

The 1630–1631 plague outbreak in Venice offers an opportunity to test these hypotheses. Advantages of this for hypothesis testing include the absence of an effective intervention (vaccines, antibiotics), restricted movement, detailed daily death records unrestricted by privacy laws, and a complete, uninterrupted epidemic cycle rarely seen in modern outbreaks.^13–15^ These features, combined with clear geographical boundaries between parishes, make the Venice outbreak an ideal “natural experiment” for testing basic generalizable principles of epidemic spread.^16^ Here, we test the hypotheses that the distribution of mortality across parishes both had increasing entropy over the course of the 1630–1631 Venetian plague, and approached a maximum entropy distribution.

## Results

### Descriptive Statistics

A total of 113,111 individuals across 50 parishes were considered at risk for infection on January 1, 1629. Among the 1629 population estimates of the 50 parishes, the median parish population was 1,734 (SD 1,802). A total of 42,538 deaths were recorded over the specified time span. The mean cumulative mortality of a parish from January 1, 1629 to December 31, 1631 was 37% (SD 10%). The progression in the geographic distribution of cumulative mortality over time is depicted in Figure 1 and Supplementary Video S1 online.

**Fig. 1.**
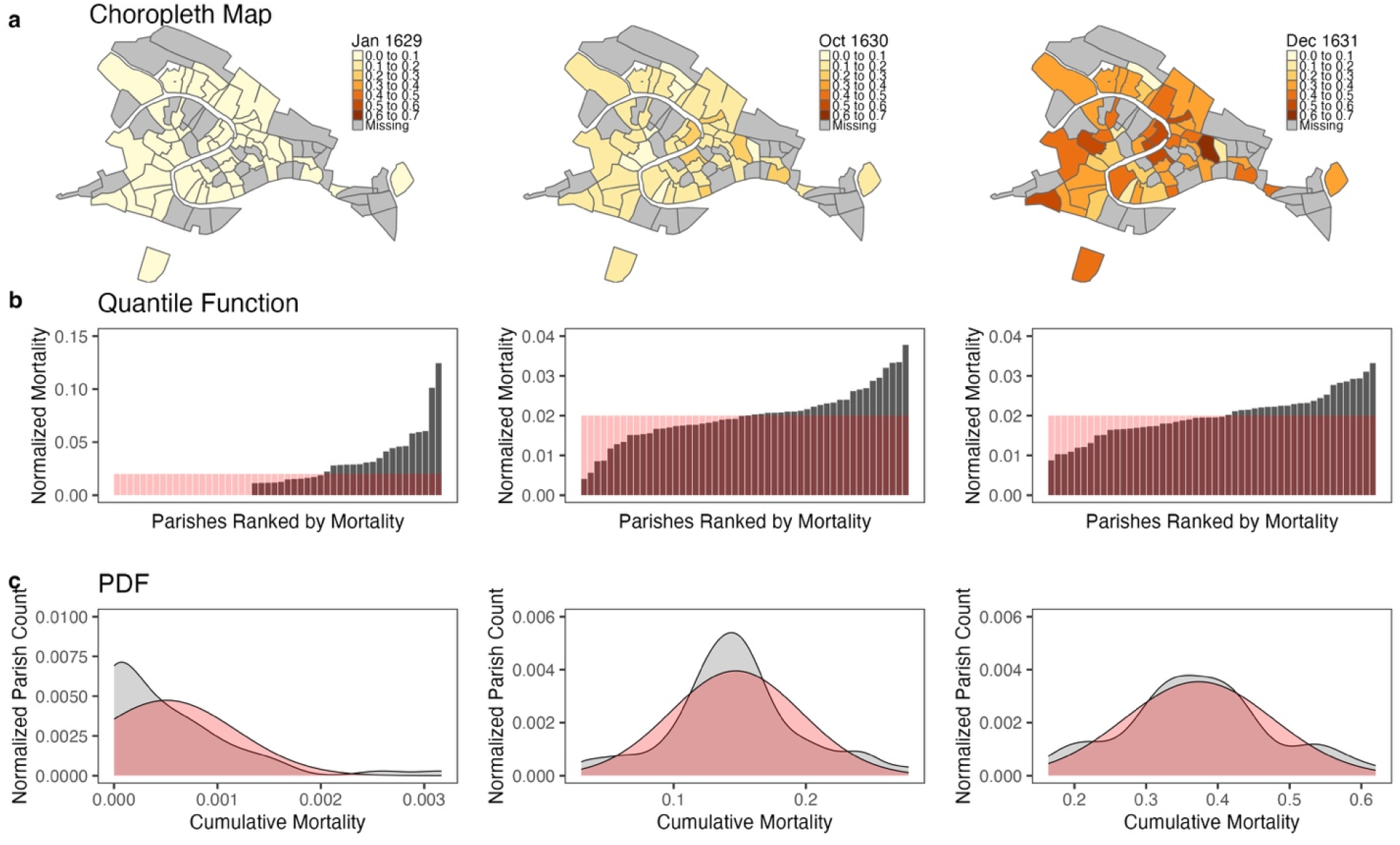
(**a**) Choropleth maps, (**b**) quantile function bar charts (ranked parishes by their normalized death rates), and (**c**) smooth probability density function (PDF) histograms (Epanechnikov kernel, *N* = 512) depicting the geographic distribution of cumulative mortality at the beginning of the epidemic (January 7, 1629), during the primary wave (October 31, 1630), and at the end of the epidemic (December 31, 1631). In (**a**), darker shades of orange correspond to higher cumulative mortalities, represented as the ratio of the total deaths in a parish to its population. The maximum entropy distribution is overlaid in red on the quantile functions in (**b**) and the PDFs in (**c**). The coefficient of variation (CV) was 1.32 (95% CI 1.00–1.65) on January 7, 1629, 0.34 (0.26–0.41) on October 31, 1630, and 0.28 (0.22–0.33) on December 31, 1631.

### Cumulative Mortality

When we analyzed the temporal progression of mortality distribution across Venice’s parishes throughout the epidemic, the quantile function of cumulative mortality (ranked parishes by their death rates) demonstrated a nearly monotonic increase in entropy, indicating a progressive shift from concentrated to dispersed mortality patterns. For each entropy value, the number of parishes with uniformly distributed mortality that would result in the same entropy value (effective number of parishes) was also evaluated. At the outbreak’s onset (January 1, 1629), the distribution showed relatively low entropy of 1.99 nats (95% confidence interval (CI) 1.20–2.53 nats), equivalent to an effective number of 7.32 parishes (3.32–12.55 parishes)— equivalent to the same entropy as an even distribution across only 7-8 parishes. By the epidemic’s later stages (December 29, 1631), entropy had increased to 3.87 nats (3.86–3.89 nats), equivalent to an effective uniform distribution across 47.9 parishes (47.5–48.9 parishes), approaching the maximum attainable 50 parishes. This transition toward uniformity was further quantified using Kullback–Leibler (KL) divergence from a discrete uniform distribution (negentropy from a uniform), a maximum entropy distribution, which decreased from 1.92 nats (1.38–2.71 nats) at the outbreak’s start to 0.038 nats (0.023–0.054 nats) by December 1631, indicating the distribution had become nearly uniform. The time-series of entropy and negentropy of the quantile function are displayed in Figure 2.

**Fig. 2.**
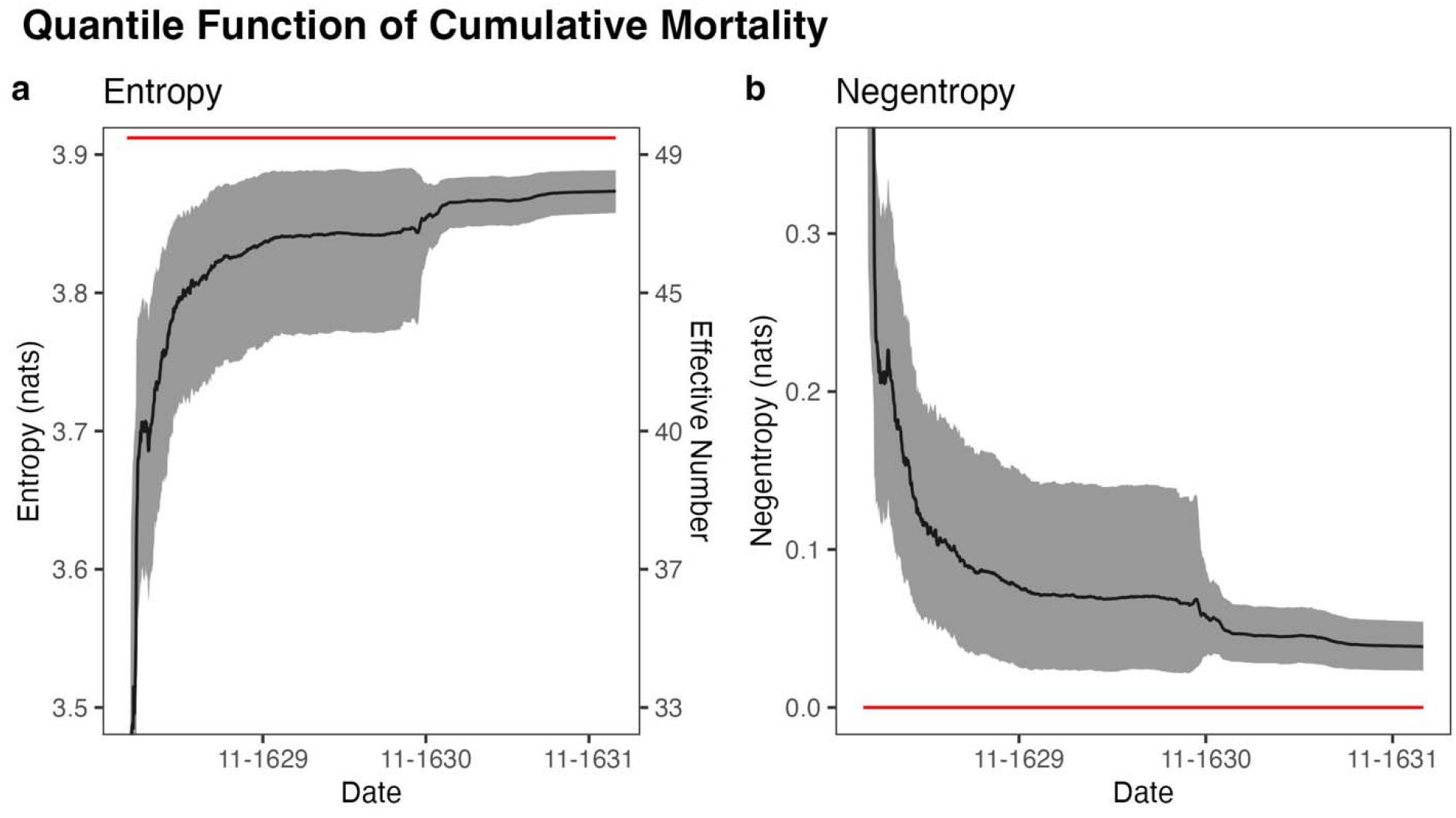
Time-series graphs for every day between January 1, 1629 and December 31, 1631 of (**a**) entropy (left axis) and effective number of parishes (right axis) of the distribution of cumulative mortality across parishes and (**b**) Kullback–Leibler divergence of the distribution of cumulative mortality across parishes from a uniform distribution. The red lines depict the maximum possible entropy and minimum possible negentropy, respectively, corresponding to an even distribution across all 50 parishes. Bootstrap 95% CIs (*N* = 999) for each day are depicted in gray.

The probability density function (PDF) of cumulative mortality (parish density in death rate bins) decreased in entropy prior to the largest wave in fall of 1630 and increased thereafter, indicating a shift in the distribution of variation in cumulative mortality toward a normal distribution after the height of the epidemic. At the outbreak’s onset, the distribution showed relatively low entropy of 0.63 nats (0.32–0.93 nats). A peak of 1.82 nats (1.55–1.89 nats) was reached on June 17, 1631. By the end of the epidemic, entropy had slightly decreased to 1.75 nats (1.53–1.87 nats). The PDF transitioned toward a Gaussian distribution, another maximum entropy distribution, over the course of the epidemic. This was quantified with Kullback–Leibler (KL) divergence from a binned Gaussian distribution with identical mean and variance (negentropy from a Gaussian). Negentropy decreased from 0.84 nats (0.71–1.42 nats) at the outbreak’s start to a minimum of 0.071 nats (0.050–0.31 nats) on May 5, 1629 before slightly increasing to 0.12 nats (0.083–0.35 nats) by its end, indicating the distribution had become relatively normal. Thus, the distribution of variation in cumulative mortality between parishes was consistent with that of a stochastic process. The time-series of entropy and negentropy of the PDF are displayed in Figure 3.

**Fig. 3.**
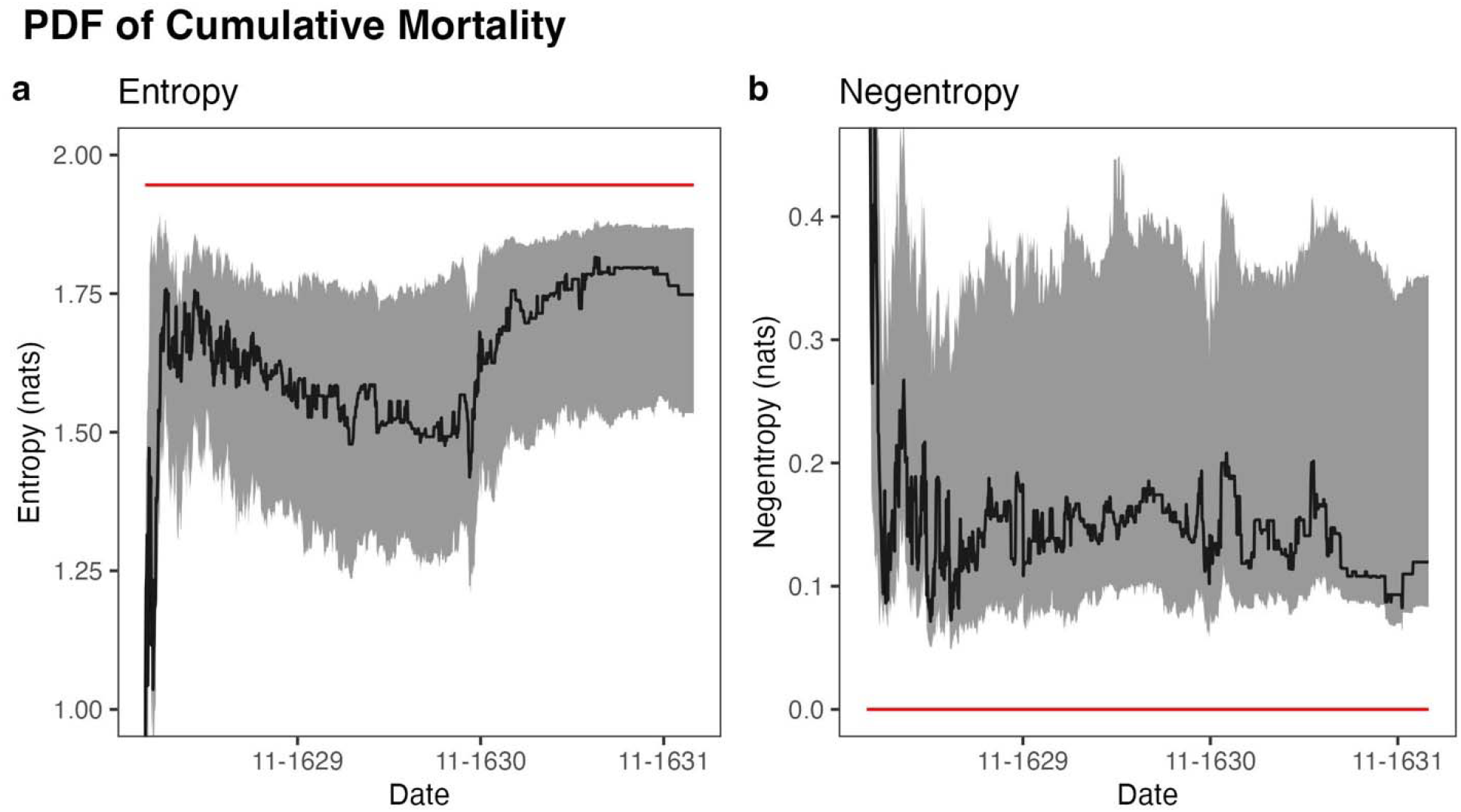
Time-series graphs for every day between January 1, 1629 and December 31, 1631 of (**a**) entropy of the binned distribution of parish density across cumulative mortalities and (**b**) Kullback–Leibler divergence of the binned distribution of parish density across cumulative mortalities from a binned Gaussian distribution with the same mean and variance as the data. The red lines depict the maximum possible entropy and minimum possible negentropy, respectively. Bootstrap 95% CIs (*N* = 999) for each day are depicted in gray.

### Instantaneous Mortality Rate (Daily)

The distribution of instantaneous mortality rate across Venice’s parishes (quantile function of instantaneous mortality) approached maximal entropy during the largest wave in the fall of 1630 and minimal entropy at the beginning and end of the epidemic, indicating that the distribution of daily deaths was only relatively uniform at the height of the epidemic. At the outbreak’s onset, the distribution showed relatively low entropy of 1.99 nats (1.23–2.52 nats). In the fall of 1630 (November 29), entropy had increased to a peak of 3.75 nats (3.68–3.81 nats). However, by the end of the epidemic, the distribution once again showed relatively low entropy of 1.96 nats (1.05–2.52 nats). The uniformity of the distribution of daily deaths was further quantified using negentropy from a uniform, which decreased from 1.92 nats (1.40–2.68 nats) at the outbreak’s start to a minimum of 0.16 nats (0.10–0.23 nats) in the fall of 1630 (November 29) before increasing to 1.95 nats (1.39–2.86 nats) by its end, indicating that the distribution was only relatively uniform at the height of the epidemic. The lack of uniformity of the distribution before and after the peak in cases does not indicate that the likelihood of infection was heterogeneous across parishes, but rather that new cases did not occur frequently enough for the uniform pattern to be observed over a single day. The time-series of entropy and negentropy of the instantaneous quantile function are displayed in Supplementary Fig. S2 online.

The distribution of parish density across instantaneous mortality rates (PDF of instantaneous mortality) approached maximal entropy during the largest wave in the fall of 1630 and minimal entropy at the beginning and end of the epidemic, indicating that the distribution of variation in new deaths was only relatively normal at the height of the epidemic. At the outbreak’s onset, the distribution showed relatively low entropy of 0.63 nats (0.32–0.95 nats). In the fall of 1630 (November 7), entropy had increased to a peak of 1.90 nats (1.72–1.92 nats). However, by the end of the epidemic, the distribution once again showed relatively low entropy of 0.65 nats (0.27–0.93 nats). The PDF transitioned toward normality as quantified by negentropy from a Gaussian, which decreased from 0.84 nats (0.71–1.43 nats) at the outbreak’s start to a minimum of 0.088 nats (0.075–0.27 nats) in the fall of 1630 (November 23) before increasing to 0.92 nats (0.79–1.56 nats) by its end, indicating that the distribution of variation in new deaths was only relatively normal at the height of the epidemic. As with the daily quantile function, this lack of normality may reflect an insufficient number of cases to observe the normal pattern that would be expected from a simple stochastic process on a daily basis. The time-series of entropy and negentropy of the instantaneous PDF are displayed in Supplementary Fig. S3 online.

### Sestieri

Venice comprises 6 districts known as sestieri, each of which contains an average of 8.33 parishes (SD 3.20 parishes). If cumulative mortality had been clustered geographically, the mean cumulative mortality of the parishes in each sestiere would vary across sestieri. However, such variation was not observed: across the 6 sestieri, the mean parish cumulative mortality rate was 38% with a standard deviation of only 1.5%. Analyzing entropy on the sestiere level provides insight on the behavior of entropy in areas with fewer subregions. The quantile function of cumulative mortality had increasing entropy and closely approximated a maximum entropy distribution within all 6 sestieri (Figure 4).

**Fig. 4.**
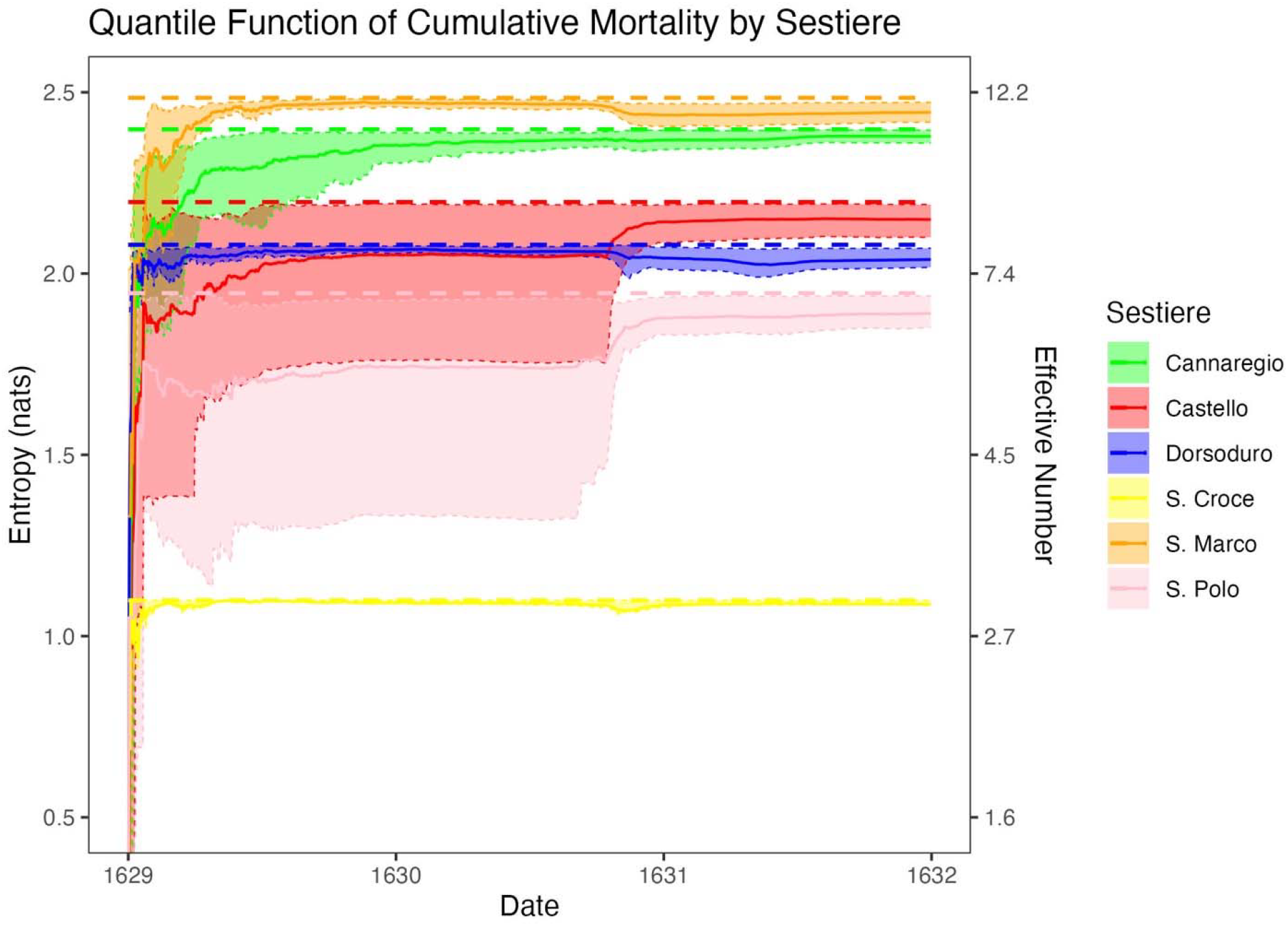
Entropy (left axis) and effective number of parishes (right axis) of the distribution of cumulative mortality across parishes for each sestiere, indicated with colors. The thick dotted lines indicate the maximum possible entropy of each sestiere, or the log of the number of parishes in that sestiere, the thin dotted lines indicate bootstrap 95% CIs (*N* = 999), and the solid lines indicate observed entropy.

## Discussion

The entropy of the geographic distribution of mortality during the 1630–1631 Venetian plague was analyzed to assess the degree to which stochastic factors influence disease spread. After the primary wave in the fall of 1630, both the quantile function (Fig. 1b) and PDF (Fig. 1c) of cumulative mortality had increasing entropy and approached a maximum entropy distribution, indicating that the spread of plague could in large part be explained by stochastic factors. This finding was robust to the choice of geographic scale, as the same behavior was observed in the subsets of parishes within each of Venice’s 6 sestieri. Additionally, at the height of the epidemic, the geographic distribution of instantaneous mortality approached a maximum entropy distribution. These findings suggest that heterogeneity in infections between regions during an epidemic may arise from random fluctuations rather than predictable factors.

The distribution of infections across communities has had increasing entropy and approached a maximum entropy distribution in outbreaks of other diseases, including SARS-CoV-2 and Ebola virus disease.^17–19^ During the COVID-19 pandemic, many models attributed variation in the cumulative prevalences in US states to heterogeneous public health practices.^6,20,21^ If this were the case, regions with high initial prevalences would have had consistently higher prevalences over the course of the epidemic. However, the distribution of cumulative prevalence across US states eventually approached a uniform distribution, indicating that this initial variation was primarily caused by stochastic factors (e.g. the timing of introduction).^19^ The distribution of infection over geographical regions in endemic diseases has also been monitored. The stochastic SIS model predicts that the PDF will approach an approximate geometric distribution when the reproduction number (*R*_*0*_) is less than one and a Gaussian distribution when *R*_*0*_ is greater than one.^22^ This has been verified in subcritical diseases such as trachoma and onchocerciasis at the village level, and trachoma and leprosy at the district level.^7,23–25^ The parameters of the maximum entropy distribution approached by the PDF of an epidemic may suffice to convey much of the epidemic’s distributional information. Additionally, for a given transmission model, it is possible to formulate a fundamental equation and equations of state relating the epidemiological interpretation of entropy to the thermodynamic definition.^11,26^ For example, for an SIS model with *R*_*0*_ > 1, the mean approaches 1-1/*R* and the variance approaches *N*/*R*.^27^ Because the entropy of the Gaussian distribution is known, it can be written in a fundamental equation as a function of the other state variables, such as *I* at equilibrium and *N*.^28^ This framework allows epidemiological parameters such as *R*_*0*_ to be estimated.

Spatial entropy has previously been used to characterize other attributes of epidemics.^9–11,26,29–31^ For instance, while this study focused on the end stages of an epidemic, a previous study used spatial entropy to predict the spatial source of epidemics.^31^ In the study, the metapopulation was represented with a directed graph and the MaxEnt method was used to reweight ensemble trajectories. However, maximum entropy distributions themselves were not used as models for the distribution of infections. The authors ran several epidemic simulations and found that their model consistently included the true infectious source among the top 5 most probable predictions. A second study used spatial entropy to identify when coronavirus infection was maximally dispersed in various countries.^9^ The authors considered non-cumulative incidence rate, and therefore used entropy to determine the time of maximum diffusion of infections. They found that all of the countries they considered reached maximal dispersion within a three-month window, and that the same methodology worked on the city-level. However, the specific maximum entropy distribution each region approached was not considered.

Limitations of this study include the consideration of deaths from all causes. While a concurrent smallpox outbreak had the potential to create a substantial difference between the observed distribution of deaths and the distribution of plague cases, particularly prior to the first wave of plague (9/1/1630–12/31/1630).^13^ However, if smallpox spread in such a way that location within the city did not matter, as was the case with the plague, this disparity would be minimal. The parish of San Marco was excluded from the dataset because it served as a placeholder for deaths from drowning and those in prison, which resulted in an early spike in cases not representative of parish transmission (see Supplementary Fig. S4–5 online).^14^ As deaths accumulated, instantaneous mortality was increasingly underestimated as parish populations were assumed to be fixed. However, because cumulative mortality was uniform across all parishes, this effect was symmetric. Finally, while entropy necessarily increases in the early stages of an epidemic due to cases occurring in regions for the first time, entropy increased at a faster rate than would be expected from this effect alone, indicating progression towards a maximum entropy distribution.

The ‘clean’ experimental conditions of the Venetian outbreak (lacking modern interventions, mass transit, or privacy restrictions) allowed us to observe fundamental patterns of disease spread that may be obscured in modern datasets. Modern outbreaks are often controlled before reaching distributional equilibrium, making historical datasets particularly valuable for testing theories about long-term epidemic behavior.^32^ Understanding these basic principles can improve our ability to predict disease distribution patterns in any epidemic, regardless of the specific pathogen or time period. For instance, the effect of interventions could be quantified with Kullback–Leibler divergence of the observed distribution of infections from the maximum entropy distribution being approached. Additionally, maximum entropy distributions could be used to benchmark more complex models or guide early outbreak response under uncertainty due to their low bias. This same entropy-based analysis could be conducted on geospatial data from any epidemic, and we hypothesize that the entropy of other epidemics will similarly increase and reach a maximum entropy distribution with sufficient time. Currently, the consistency of this pattern across epidemics of varying diseases and populations is promising, as it suggests a reliable method for predicting the distribution of cases across communities using maximum entropy distributions. The findings of this study therefore indicate that before complex models are developed to explain observed heterogeneity in the distribution of infections across areas, the degree to which this heterogeneity could be explained by chance alone merits consideration.

## Methods

### Data collection and cleaning

Venetian census data of parish populations from both 1586 and 1696 was linearly interpolated to obtain estimates of parish populations in 1629 (see Supplementary Fig. S6 online).^33^ Death counts in each of 50 parishes recorded daily between January 1, 1629 and December 31, 1631 in Venetian necrologies were normalized by parish populations to calculate cumulative mortality (see Supplementary Fig. S7 online). Because all-cause deaths, rather than infections, were recorded, all-cause deaths were used to approximate infections. However, all-cause deaths closely match plague-related deaths because plague was the primary cause of death during the time.^15^ Cumulative mortality was used to avoid an upper bound on entropy values due to insufficient cases. However, because cumulative mortality necessarily increases over the course of an epidemic, we used intensive entropy, or the average entropy per case. Boltzmann-Gibbs-Shannon entropy in nats (defined as – 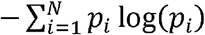, where *p*_*i*_ is the probability in bin *i*) was used to relate our findings to the number of combinatorial arrangements of cases across regions that would result in the observed distribution. The effective number of parishes, or the number of evenly distributed parishes that would result in the same entropy value, was evaluated as the exponential function of entropy.^34^ Because parish names were inconsistent between censuses and death records, manual matching was occasionally required. 20 parishes were excluded from the initial dataset containing 70: one parish (S. Marcilian) was not recorded in the 1696 census data, 18 had no data in the primary outbreak period (9/1/1630–12/31/1630), and one (S. Marco) was a placeholder for deaths from drowning, deaths in prison, and deaths with other ambiguous locations due to its role as an administrative district.^14^ For included parishes, missing data points were replaced with zeroes.

### Data analysis and modeling

The geographic distribution of mortality was represented with both a quantile function and a probability density function (PDF). The quantile function was the bar chart of parishes ordered by mortality while the PDF was approximated by a histogram of parish counts binned by mortality under Sturges’s Rule.^35^ Kullback–Leibler (KL) divergence from maximum entropy distributions, or negentropy, was used to measure the extent to which the distribution of mortality approximated a maximum entropy distribution. KL divergence can be interpreted as the expectation of the logarithmic difference between the observed probability distribution *P* and the model distribution *Q*, defined as 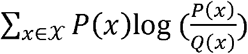. For the quantile function, KL divergence from a discrete uniform distribution was calculated. For the PDF, KL divergence from a binned Gaussian distribution with identical mean and variance to the data was also calculated.^36^

Bootstrap 95% CIs for both entropy and KL divergence were calculated by resampling parishes (*N* = 999) and evaluating the 2.5% and 97.5% intervals at each time point to test for bias in parish selection. Entropy and KL divergence were also calculated and bootstrapped within each sestiere to identify potential proximity-based heterogeneity in this pattern. The geographic distribution of cumulative mortality for every month in the specified time period was visualized with choropleth maps, the base code for which was sourced from Lazzari et al.^15^ The entropy of daily mortality and the entropy when the parish of S. Marco was included were calculated as sensitivity analyses. All analyses were conducted using R (Version 4.2.0 for MacOS).^37^

## Data Availability

The dataset analyzed during the current study is available in the Venice-plague-epidemic-paper repository created by Lazzari et al., https://github.com/ggrrll/Venice-plague-epidemic-paper.

## Acknowledgements

We would like to thank Dr. Gianrocco Lazzari, Dr. Giovanni Colavizza, and their collaborators for compiling and digitizing the database in a previous study. The work presented in this article is funded by the National Eye Institute (NEI) at the National Institutes of Health (NIH) (grant numbers R01EY025350 and R01EY024608). This work was supported in part by a grant from an unrestricted grant from Research to Prevent Blindness (PI: JLD).

## Author contributions

All authors participated in the design of the experiment. J.L.H. performed the analysis. J.L.H. and T.M.L. wrote the initial draft. All authors reviewed and approved the manuscript.

