## Supplementary Information for "Spread of the plague in Venice, 1630–1631: epidemic entropy in a “natural experiment”"

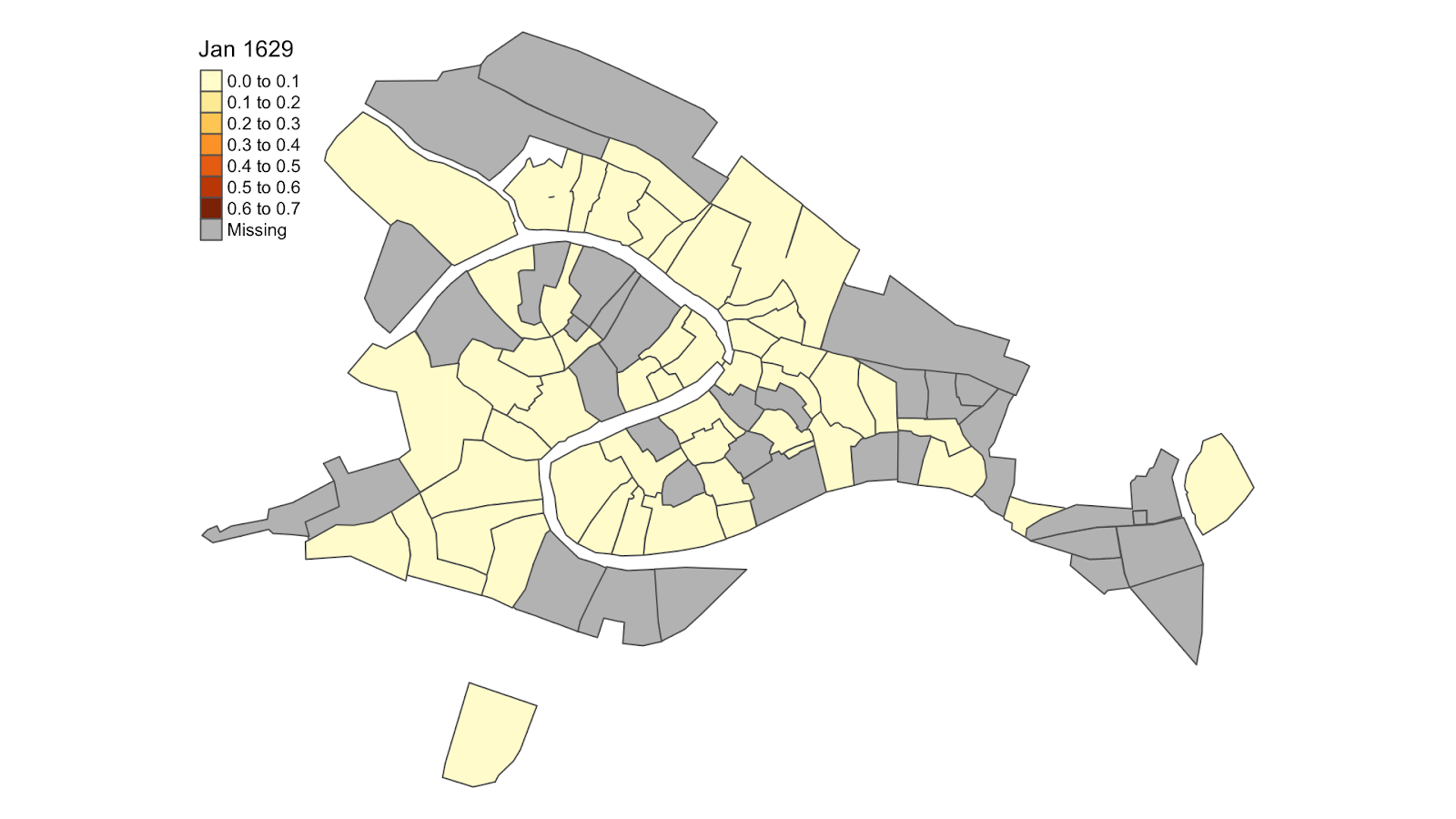

Supplementary Video S1. A series of choropleth maps depicting the geographical distribution of cumulative mortality in every month between January 1, 1629 and December 1, 1631. Darker shades of red represent higher cumulative mortalities; see the legend for more details.

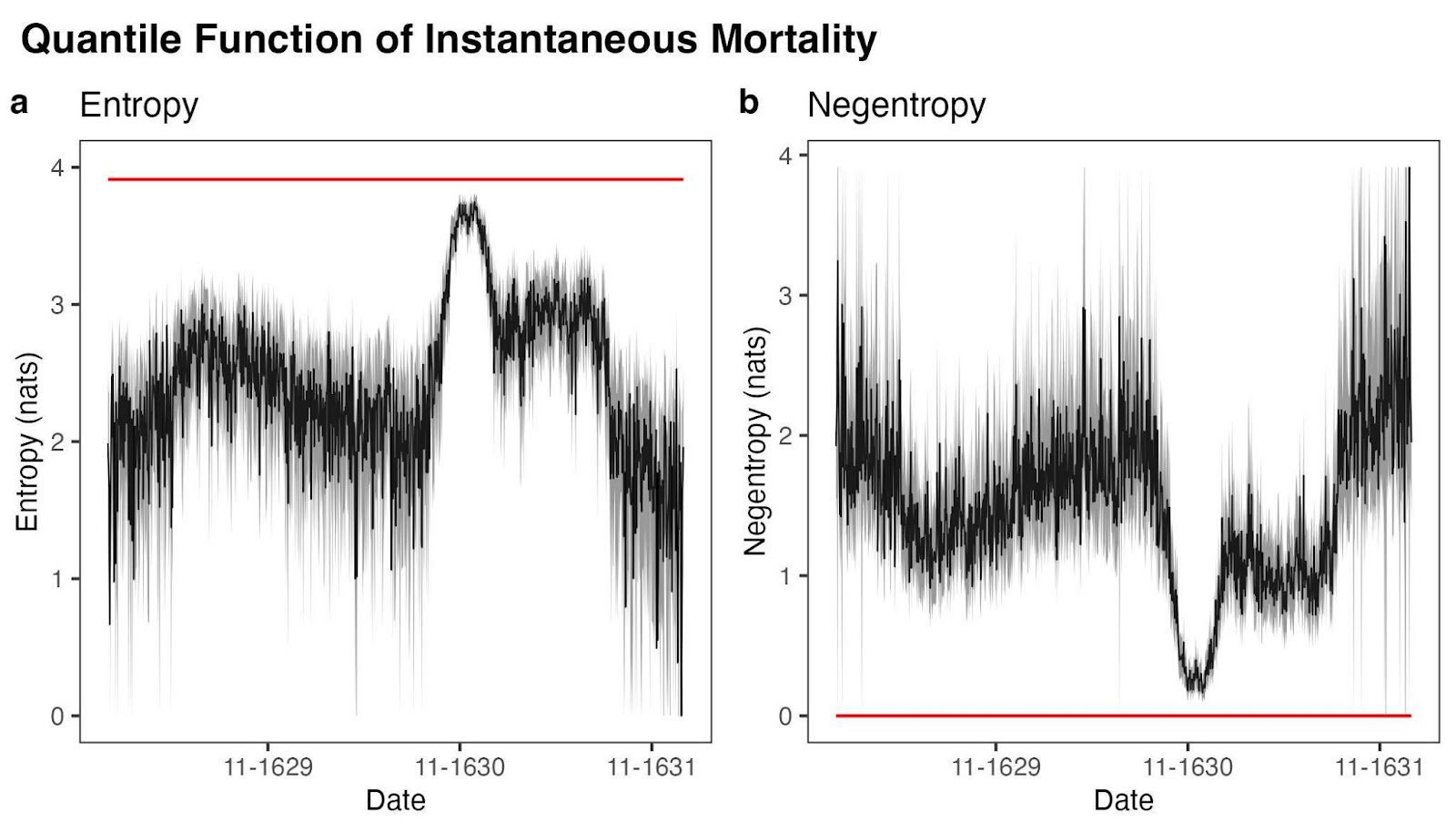

Supplementary Figure S2. Time-series graphs for every day between January 1, 1629 and December 31, 1631 of (**a**) entropy of the distribution of daily mortality across parishes and (**b**) Kullback–Leibler divergence of the distribution of daily mortality across parishes from a uniform distribution. The red lines depict the maximum possible entropy and minimum possible negentropy, respectively. Bootstrap 95% CIs (*N* = 999) for each day are depicted in gray.

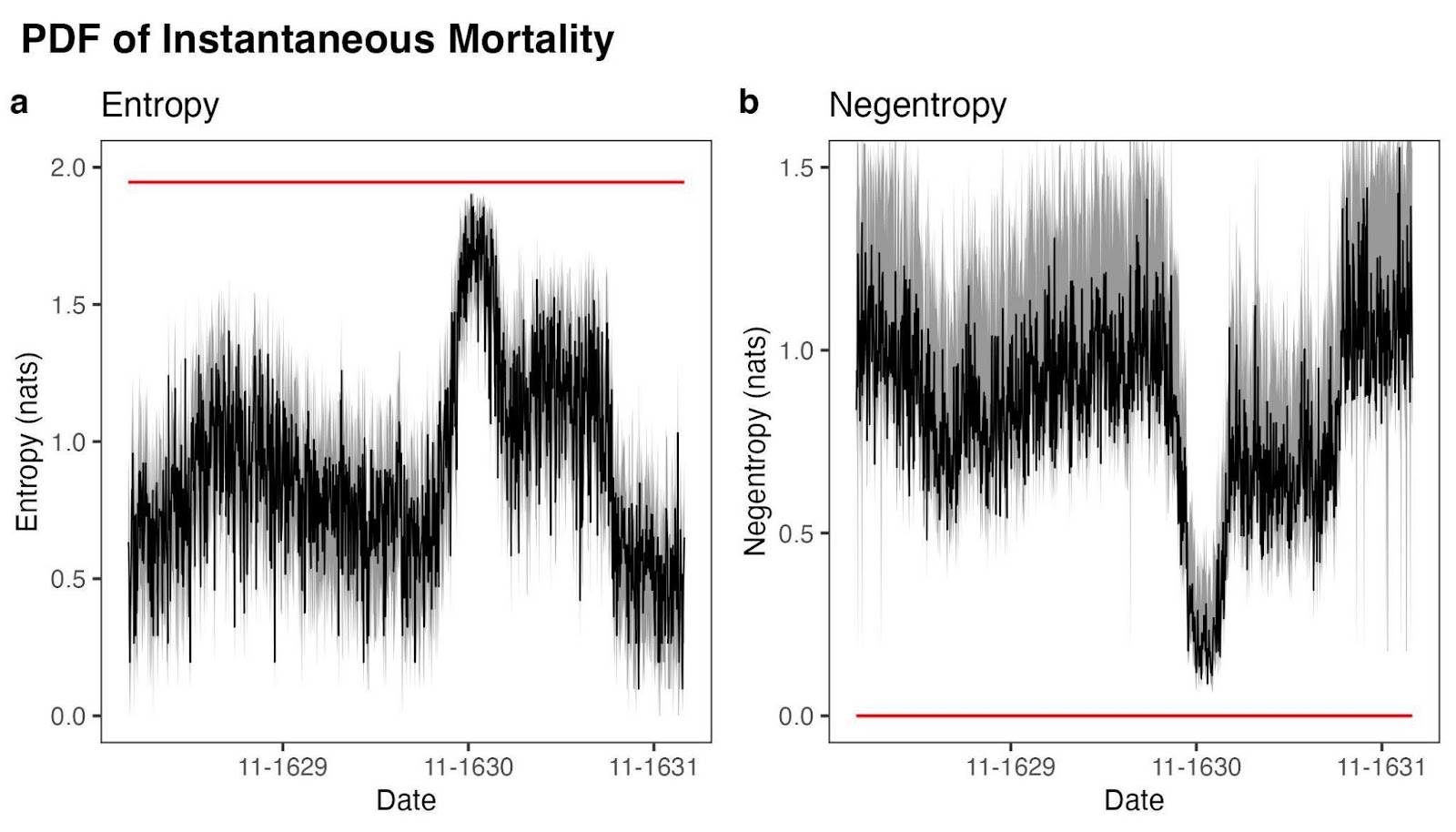

Supplementary Figure S3. Time-series graphs for every day between January 1, 1629 and December 31, 1631 of (**a**) entropy of the binned distribution of parish density across different daily mortalities and (**b**) Kullback–Leibler divergence of the binned distribution of parish density across different daily mortalities from a binned Gaussian distribution with the same mean and variance as the data. The red lines depict the maximum possible entropy and minimum possible negentropy, respectively. Bootstrap 95% CIs (*N* = 999) for each day are depicted in gray.

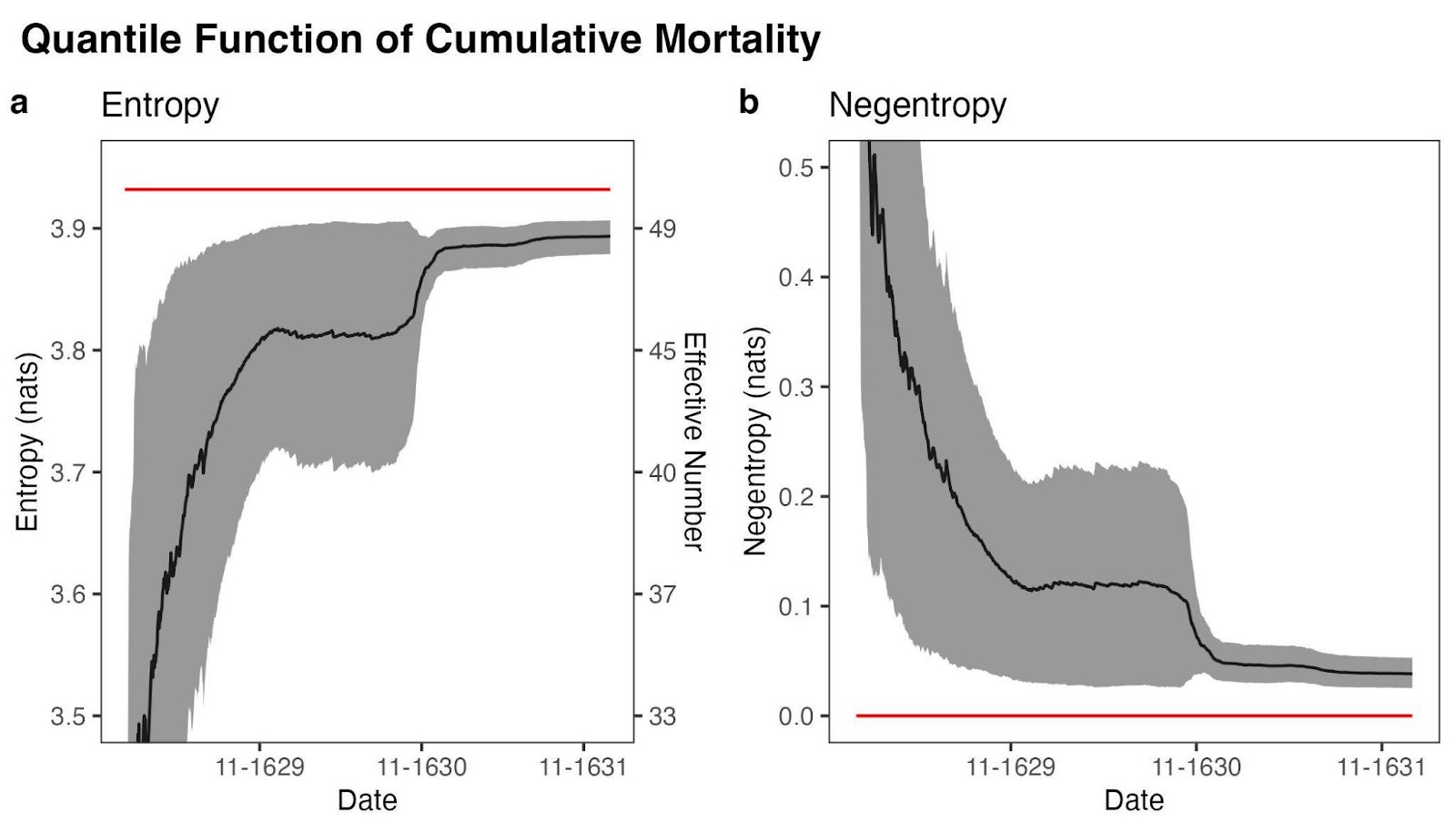

Supplementary Figure S4. Time-series graphs for every day between January 1, 1629 and December 31, 1631 of (**a**) entropy (left axis) and effective number of parishes (right axis) of the distribution of cumulative mortality across parishes and (**b**) Kullback–Leibler divergence of the distribution of cumulative mortality across parishes from a uniform distribution when S. Marco is included. The red lines depict the maximum possible entropy and minimum possible negentropy, respectively. Bootstrap 95% CIs (*N* = 999) for each day are depicted in gray.

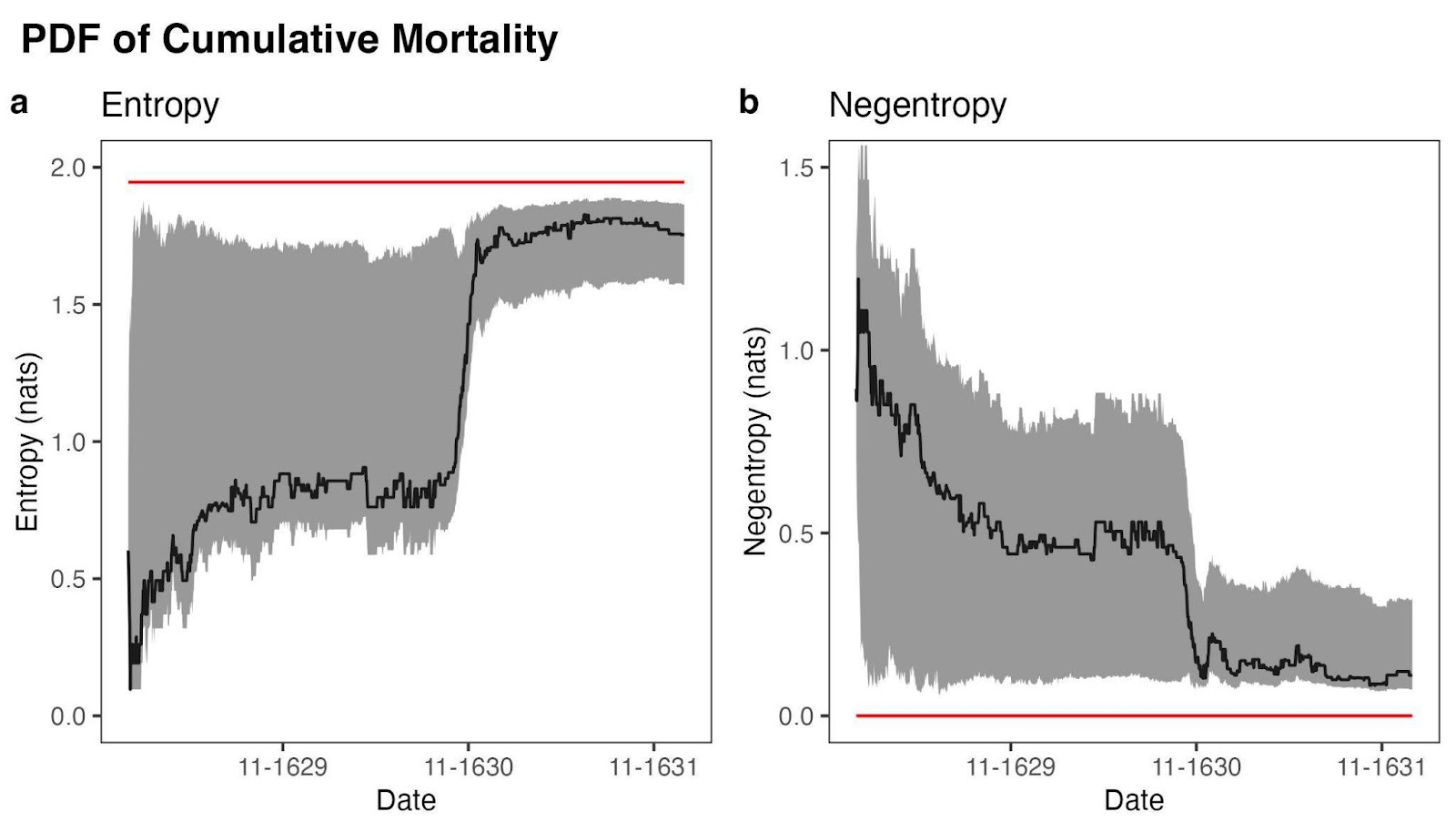

Supplementary Figure S5. Time-series graphs for every day between January 1, 1629 and December 31, 1631 of (**a**) entropy of the binned distribution of parish density across different cumulative mortalities and (**b**) Kullback–Leibler divergence of the binned distribution of parish density across different cumulative mortalities from a binned Gaussian distribution with the same mean and variance as the data when S. Marco is included. The red lines depict the maximum possible entropy and minimum possible negentropy, respectively. Bootstrap 95% CIs (*N* = 999) for each day are depicted in gray.

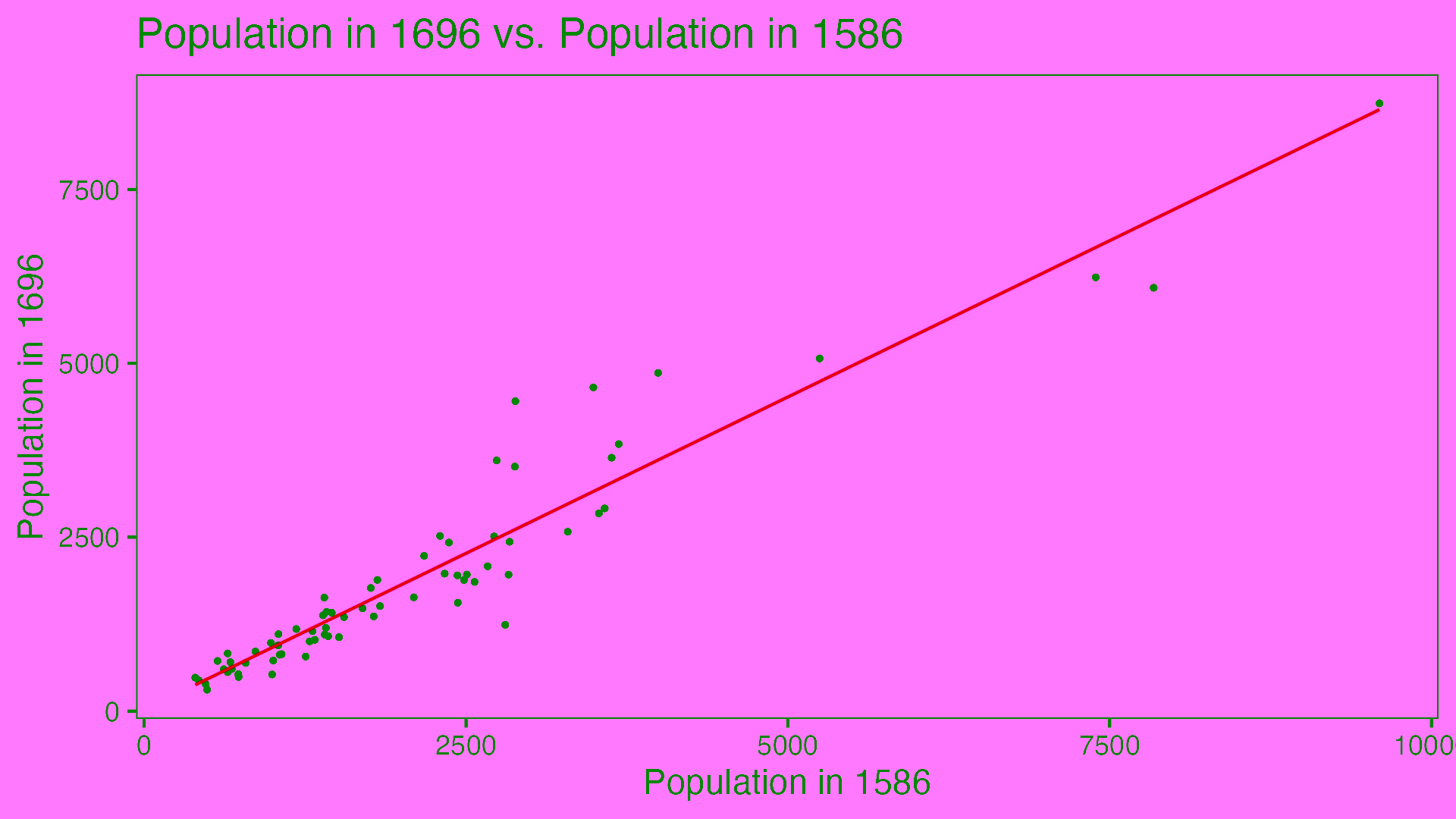

Supplementary Figure S6. Linear model relating population in 1696 (y-axis) to population in 1586 (x-axis). *r*^2^ = 0.952.

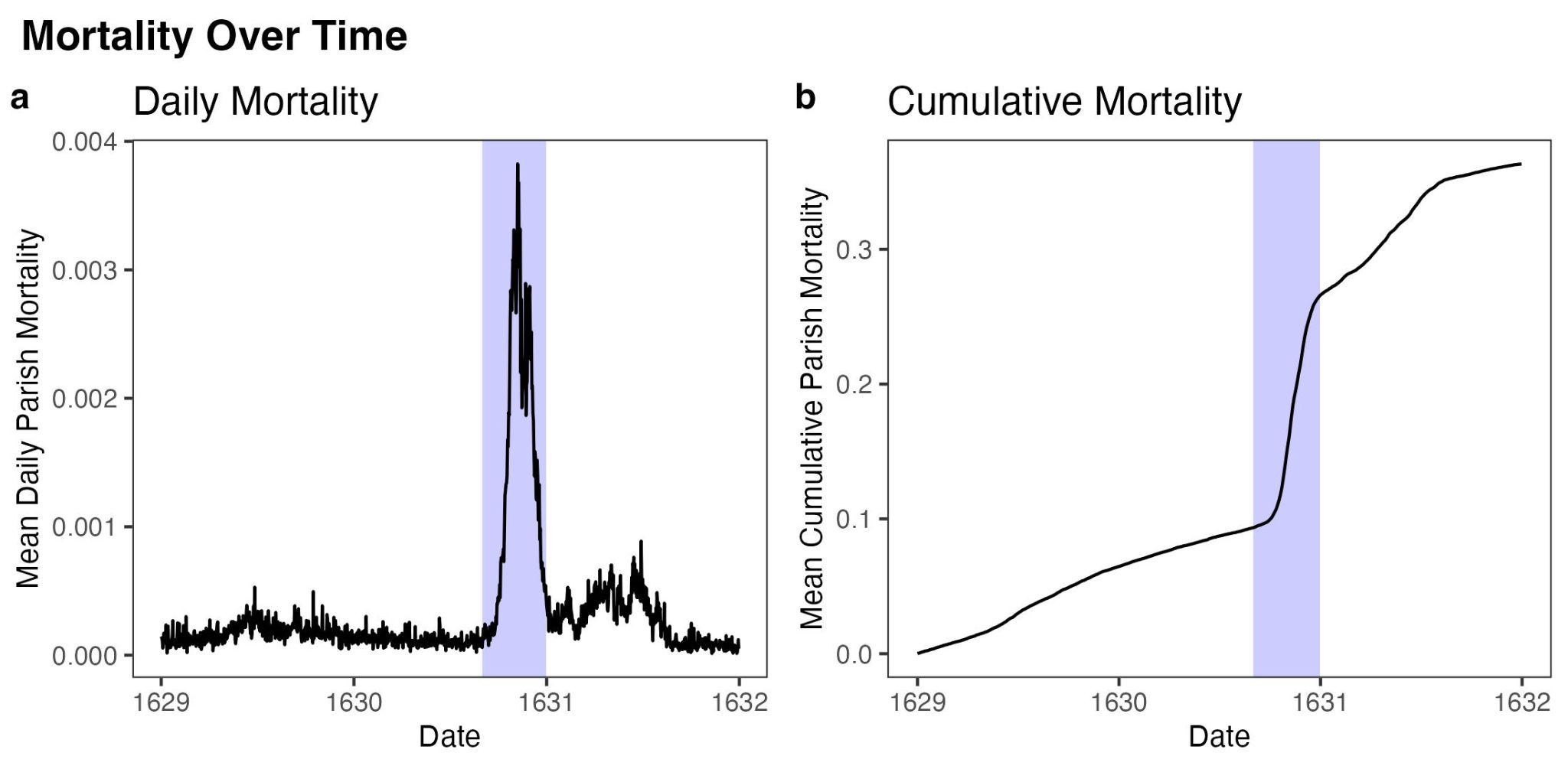

Supplementary Figure S7. Time-series graphs for every day between January 1, 1629 and December 31, 1631 of (**a**) mean daily mortality across parishes and (**b**) mean cumulative mortality across parishes. The blue window depicts the primary outbreak window from September 1, 1630 to December 31, 1630.
